# Site-variable allocation ratios in randomized controlled trials: implications for sample size, recruitment efficiency, and statistical analysis

**DOI:** 10.1101/2024.11.03.24316666

**Authors:** Pavel S. Roshanov

**Affiliations:** Department of Medicine (Division of Nephrology), Western University, London, ON; Department of Epidemiology & Biostatistics, Western University, London, ON; Population Health Research Institute, Hamilton, ON

## Abstract

**Introduction:** In multicentre randomized trials, some sites face logistical constraints that specifically affect their ability to recruit into one arm of the trial more than other arms. Often these are greater limits on their ability to deliver one of the study interventions. This paper proposes the use of allocation ratios that differ by site to increase recruitment capacity in asymmetrically constrained sites.

**Methods:** Simulations of randomized trials assessed the impact of several allocation ratios (1:1 to 1:5)—and variation of ratios across sites—on sample size and recruitment capacity, and evaluated several adjustment approaches for time-to-event, binary, and continuous outcomes to prevent bias from site-variable allocation ratios.

**Results:** Deviating from 1:1 allocation increases recruitment capacity within sites facing asymmetric constraints faster than it increases sample size requirements. For instance, a 1:3 ratio increased sample size by 35% but doubled the hypothetical recruitment capacity with fewer sites. The bias in treatment effect estimates that occurs when the baseline risk or outcome mean differ between sites allocated with different ratios was readily prevented with simple covariate adjustment or stratification by site or allocation ratio.

**Conclusions:** Site-variable allocation ratios may relieve recruitment bottlenecks caused by asymmetric constraints in trial procedures that affect some of the sites in a trial. Accounting for the variation in allocation ratios during analysis is necessary to ensure unbiased treatment effect estimates. This strategy is particularly relevant for trials with low marginal costs for participant recruitment and follow-up, such as many large pragmatic trials embedded in routine care.

## 1.0 INTRODUCTION

In multicentre randomized trials, some study sites encounter logistical hurdles that disproportionately affect their capacity to enroll participants into one arm of the trial compared to others. This constrains their recruitment capacity.

Consider a remote monitoring and virtual care program for people discharged home after major surgery. The program was evaluated in a trial where individual patients were randomly allocated at the time of hospital discharge, in a 1:1 ratio, to continue with usual post-discharge care or to receive post-discharge virtual care by dedicated healthcare team at each study site.[1] This intervention involved frequent video assessments by nurses and physicians. Many patients were eligible and most were eager to participate in the trial, but recruitment at each site was limited by the number of patients that the local intervention team could accommodate, and this differed across sites. The trial was conducted in the early months of the COVID-19 pandemic. With the limited nursing resources that the hospitals could contribute to the intervention under constant threat of being withdrawn, the trial needed to recruit as quickly as possible.

Consider a second example: a large pragmatic randomized trial being designed to compare the major health effects of two kinds of dialyzers (i.e., hemodialysis filters) available in routine care. Most patients are eligible for the trial and would participate. Study outcomes are captured from administrative healthcare databases. The marginal cost of enrolling and following a patient in this trial is very low and trial is planned to be large, but the investigators want to contain its recruitment to one country.

Like in the remote monitoring trial, there is an asymmetric constraint: a limitation on the number of patients that can be treated with one of the two types of dialyzers being compared. The procurement and use of medical consumables like dialyzers is often governed by contractual agreements between healthcare providers and suppliers. The agreements are designed to optimize costs and ensure consistent supply, typically through volume-based pricing structures. Healthcare organizations commit to purchasing a certain volume of products to access more favorable pricing tiers. These contracts include clauses that require a specific percentage of total usage to be of “on-contract” products to maintain the negotiated prices. For example, a hospital might agree to use a particular type of dialyzer for at least 80% of their hemodialysis procedures to secure a significant discount. As a result, some sites (dialysis units) are limited by the number of patients they can treat with one of the dialyzer before breaching their purchasing contract, but have no such constraint on the use of the comparator dialyzer (which they have on contract). Thus, the capacity to deliver one arm of the trial limits the number of patients sites can enrol in the trial. For other sites, this constraint has been removed.

### Varying allocation ratios offer a solution

Most randomized trials comparing two alternative treatments allocate patients in a 1:1 ratio, and this is usually statistically optimal (i.e., requires the fewest participants to achieve the same statistical power).[2] However, a more efficient strategy overall may be to accommodate site constraints by allowing the allocation ratio between intervention and control groups to vary by study site. This strategy can maximize recruitment capacity within the confines of intervention delivery limitations while maintaining overall statistical power and reducing the time and number of sites required to execute the trial. Because balanced allocation is still more statistically efficient, study sites that are not constrained in intervention delivery can use balanced allocation while those that are constrained can use an allocation ratio that accommodates their asymmetric constraints.

However, site selection into different allocation ratios is not random and may be associated with the outcomes under study. Therefore, the analysis requires additional adjustments to avoid bias.

This paper provides two things. The first is an exploration of the effects of different fixed and variable allocation scenarios on sample size requirements and hypothetical recruitment capacity when recruitment is constrained by asymmetric limitations in study procedures (e.g., one intervention more constrained than the other). The second is an exploration of the basic analytical adjustments required in the analysis of time-to-event, binary, and continuous outcomes to account for bias that may arise due to site selection into different allocation ratios.

## 2.0 METHODS

### 2.1 Impact of allocation ratio on sample size and recruitment capacity

This illustrative analysis was performed on simulated data. Five different intervention-to-control allocation ratio scenarios were examined: 1:1, 1:2, 1:3, 1:4, and 1:5. To calculate sample size, simulations were performed of 1000 hypothetical multicentre randomized controlled trials where each study site can only deliver the intervention to 10 patients. All sites were assumed to use the same allocation ratio for each scenario. The trial had a target power of 90% to detect a hazard ratio for death of 0.85 (i.e., a 15% relative reduction in the hazard of death) at two-sided alpha of 0.05 using the log rank test. Based on the Lakatos normal approximation[3], other assumptions made for the calculation were: 1) death rate in the in the control group is 17.0 deaths per 100 patient-years of follow-up (from a pragmatic trial in hemodialysis)[4]; 2) recruitment is planned to take 2 years; 3) the trial will be completed within 5 years (from enrollment of the first patient to termination of intervention period), and 4) 2% dropout per year.

From this followed simple calculations of the number of sites required, increase in enrolment capacity per site, and increase in sample size requirements across allocation ratios compared to 1:1.

The next exploration was of the combination of 1:1 and 1:3 allocation across sites in the same trial examined the effect of different relative contributions of each allocation ratio on power and sample size.

The last exploration in this section addressed how constant and variable allocation ratios compare in the number of study sites required. This is done using a scenario where investigators of the dialyzer trial initially recruit 48 study sites that are unconstrained in their ability to deliver the intervention but have not yet started recruiting. The investigators must decide between a uniform 1:1 allocation for all sites including any future sites (all of which would have constraints in intervention delivery), a uniform 3:5 allocation across sites (including for all unconstrained sites recruited to date), and a variable 1:1 and 1:3 allocation depending on site constraints.

### 2.2 Analysis of outcomes when allocation ratios vary by study site

The next set of analyses concerned the bias that might result from site selection into different allocation ratios.

Data were simulated for 1000 hypothetical trials. Each trial involved 120 sites, equally divided into two groups: 60 sites used a 1:1 treatment-to-control allocation ratio, and the remaining 60 sites used a 1:3 ratio. Each site enrolled 40 participants, resulting in a total sample size of 4800 participants in each simulation. Randomization was stratified by study site. Each trial had a time-to-event outcome, a binary outcome, and a continuous outcome.

All treatment effects were set to null. For the time-to-event outcome, baseline hazards were set at 0.1 for sites with a 1:1 allocation ratio and increased by 50% to 0.15 for sites with a 1:3 ratio to simulate prognostic differences that could be found between sites using different allocation ratios. Event times were simulated using exponential distributions, and random censoring times were uniformly distributed between 0 and 5 units of time, with observed times defined as the minimum of the event and censoring times. For the binary outcome, the baseline probability was established at 0.2 for 1:1 sites and increased by 50% to 0.3 for 1:3 sites, with outcomes generated using logistic models. Continuous outcomes were modeled with a baseline mean of 100 units for 1:1 sites and increased by 10 units for 1:3 sites to reflect site differences, and these outcomes were generated from normal distributions with a standard deviation of 20 units.

#### 2.2.1. Analysis Methods

These analyses used Cox proportional hazards models for time to (first) event outcomes, logistic regression for binary outcomes, and linear regression models for continuous outcomes. They included unadjusted models (with a treatment allocation indicator as the only independent variable), models adjusting for site or allocation ratio as covariates, and analyses stratified site or allocation ratio and combined using fixed-effects inverse-variance meta-analysis. The binary outcomes analysis stratified by study site used conditional logistic regression. Finally, there were random effects (intercept) models for both binary and continuous outcomes and the equivalent for the Cox model of time-to-first event data (shared frailty) for study site. Four key metrics were evaluated: the mean estimated treatment effect across simulations; mean standard error; and 95% confidence interval coverage (i.e., percentage of simulations where the confidence included the true effect).

All analyses were performed using R 4.4.1 (R Core Team [2024]).

## 3.0 RESULTS

### 3.1 Impact of allocation ratio on sample size and recruitment capacity

At an allocation ratio of 1:1, 3550 patients in total are required to meet the power requirements for the trial. Assuming that sites can recruit only 20 patients on average (limited by 10 in the intervention group), the trial needs to activate 178 sites. **Table 1** shows how recruitment capacity within sites increases more than the required sample size as the allocation ratio deviates further from 1:1. Consequently, the number of required study sites also decreases. At a 1:3 ratio, 4800 patients are required (an increase of 35%) with 1200 in the intervention group and 3600 in the control group but recruitment capacity per site triples and the number of sites required decreases to 120; 48% more sites would be required with these constraints in 1:1.

**Table 1:** Sites and recruitment capacity for different allocation ratios.

| **Allocation Ratio** | **Total Sample Size** | **Sample Size Multiplier** | **Intervention Patients** | **Control Patients** | **Mean Recruitment Capacity per Site (intervention:control)** | **Recruitment Capacity Multiplier** | **Sites Needed** |
| --- | --- | --- | --- | --- | --- | --- | --- |
| 1:1 | 3550 | 1.00 (ref) | 1775 | 1775 | 20 (10:10) | 1.00 (ref) | 178 |
| 3:5 | 3800 | 1.07 | 1425 | 2325 | 26.7 (10:16.6) | 1.33 | 143 |
| 1:2 | 4000 | 1.13 | 1400 | 2800 | 30 (10:20) | 1.50 | 133 |
| 1:3 | 4800 | 1.35 | 1200 | 3600 | 40 (10:30) | 2.00 | 120 |
| 1:4 | 5600 | 1.58 | 1080 | 4320 | 50 (10:40) | 2.50 | 108 |
| 1:5 | 6450 | 1.82 | 1290 | 5160 | 60 (10:50) | 3.00 | 108 |

Footnote: Sample size estimates are based on mean of 1000 simulated trials comparing two interventions with target 90% power to detect in a time-to-first-event analysis a hazard ratio 0.85, assuming control event rate of 17 events per 100 patient-years, 2 years of recruitment, total trial duration 5 years, and 2% dropout per year. Conditions were identical across all allocation ratio scenarios. Total sample size requirements were rounded to the nearest 25 patients.

**Table 2** illustrates the effect on power and sample size for various combinations of 1:1 and 1:3 allocation ratios for the dialyzer trial. As more patients are enrolled in a 1:1 ratio, power increases or the required sample size decreases while maintaining 90% power.

**Table 2.** Effect of allocation ratio mix on power and sample size.

| **% 1:1 ratio** | **% 1:3 ratio** | **Overall ratio**<br>**(mixed)** | **% in the**<br>**intervention**<br>**group** | **Power if n=4800** | **Sample size for 90%**<br>**power** |
| --- | --- | --- | --- | --- | --- |
| 0 | 100 | 1:3 | 25 | 90.2 | 4800 |
| 10 | 90 | 8:21 | 27.5 | 91.9 | 4500 |
| 20 | 80 | 3:7 | 30 | 93.1 | 4250 |
| 30 | 70 | 13:27 | 32.5 | 94.1 | 4075 |
| 40 | 60 | 7:13 | 35 | 94.7 | 3925 |
| 50 | 50 | 3:5 | 37.5 | 95.4 | 3800 |

Footnote: Hypothetical trial of a dialyzer with two different intervention:control allocation ratios, aiming to detect in a time-to-first-event analysis a hazard ratio 0.85, assuming control event rate of 17 events per 100 patient-years, 2 years of recruitment, total trial duration 5 years, and 2% dropout per year. Results are from 1000 simulated trials in each scenario. A sample size of 4800 patients (assuming a constant 1:3 ratio) is calculated to provide 90% power. The trial will require 120 sites if each site can enrol a fixed number of 40 patients. If, however, some of the sites commit to delivering the intervention to 20 patients, then they would use the 1:1 allocation for their 40 patients. This contributes more power because 1:1 allocation is more statistically efficient than 1:3 allocation. If 50% of the patients are allocated in a 1:1 ratio and the other 50% are allocated in a 1:3 ratio, 4800 patients provide 95.4% power to detect the effect of interest under the sample conditions. Alternatively, researchers may reduce target enrolment as more patients are enrolled in a 1:1 ratio.

How do constant and variable allocation ratios compare in the number of study sites required? Although the required sample size would be the same (assuming the same event), using the same unequal ratio leaves some underutilized capacity at sites constrained in their ability to deliver the intervention. **Table 3** illustrates the effect of uniform 1:1 ratio, uniform 3:5 (the mean of a 1:1 and 1:3 ratio), and variable 1:1 and 1:3 ratio when investigators of the dialyzer trial initially only recruit 48 unconstrained sites, of which each can randomize 40 patients in a 1:1 ratio. All other potential sites are constrained by dialyzer contracts and, although they can theoretically recruit up to 40 patients, they can only deliver the intervention to 10 patients each so 1:1 randomization would leave them contributing just 20 patients each.

**Table 3.** Effect of allocation ratio approach on the number of study sites.

| **Allocation Ratio** | **Total Patients**<br>**Required for**<br>**90% power** | **Unconstrained Sites**<br>**(fixed)** | **Constrained Sites** | **Total Required Sites** |
| --- | --- | --- | --- | --- |
| Uniform 1:1 | 3550 | 48 | 82 | 130 |
| Uniform 3:5 | 3800 | 48 | 71 | 119 |
| Variable 1:1 and 1:3 | 3800 | 48 (1:1 ratio) | 47 (1:3 ratio) | 95 |

Footnote: Hypothetical scenario where investigators of the dialyzer trial initially recruit 48 sites that are unconstrained in their ability to deliver the intervention; each can randomize 40 patients in a 1:1 ratio. Investigators now must decide how to allocate patients to minimize the number of additional sites required, recognizing that all additional sites will be constrained in their ability to deliver the intervention. Each constrained site can only deliver the intervention to 10 patients but can theoretically enrol up to 40 patients (i.e., up to 30 to control) if enabled to do so by its assigned allocation ratio.

If investigators continue with 1:1 allocation in all sites, the sample size requirement is 3550 patients, but they need 82 of the constrained sites to join the trial in addition to the 48 unconstrained sites. If they instead allocate patients in 3:5 ratio in all sites, the sample size requirement increases to 3800 patients. Unconstrained sites continue to contribute 40 patients each (15 to the intervention; 25 to control), and the trial now needs to add at least 71 more constrained sites, with each now contributing on average 27 patients (10 to intervention, 17 to control). Finally, the variable scenario requires just 47 constrained sites to be added to the trial because it allows the constrained sites to reach maximal recruitment efficiency and contribute 40 patients each at 1:3 allocation ratio.

### 3.2 Analysis of outcomes when allocation ratios vary by study site

**Tables 4** summarize the results of different analysis methods across time-to-event, binary, and continuous outcomes. For all three outcome types, failing to account for at least one of allocation ratio or study site led to a biased estimate of treatment effect (which was introduced by design in the simulation). Covariate adjustment or stratification for study site or allocation ratio eliminated bias for all outcome types and assured appropriate confidence interval coverage. Shared frailty/random effects models only partly reduced bias.

**Table 4.** Results of analysis evaluating different methods of adjustment for variable allocation.

| **Time-to-event outcome<sup>†</sup>** | **Mean Hazard Ratio** | **Mean log SE** | **95% CI Coverage (%)** |
| --- | --- | --- | --- |
| No Adjustments | 0.898 | 0.0598 | 56.5 |
| Site as Covariate | 1.00 | 0.0623 | 94.8 |
| Allocation Ratio as Covariate | 1.00 | 0.0619 | 95.2 |
| Stratified by Site | 1.00 | 0.0630 | 95.1 |
| Stratified by Allocation Ratio | 1.00 | 0.0619 | 95.2 |
| Shared Frailty by Site | 0.921 | 0.0604 | 72.0 |
| **Binary outcome<sup>‡</sup>** | **Mean Odds Ratio** | **Mean log SE** | **95% CI Coverage (%)** |
| No Adjustment | 0.866 | 0.0696 | 45.2 |
| Site as Covariate | 1.00 | 0.0734 | 94.4 |
| Allocation Ratio as Covariate | 1.00 | 0.0725 | 94.7 |
| Stratified by Site | 1.00 | 0.0725 | 94.7 |
| Stratified by Allocation Ratio | 1.00 | 0.0725 | 94.7 |
| Random Effect for Site | 0.904 | 0.0717 | 71.0 |
| **Continuous outcome<sup>§</sup>** | **Mean difference** | **Mean SE** | **95% CI Coverage (%)** |
| No Adjustment | -2.67 | 0.613 | 0.7 |
| Site as Covariate | -0.006 | 0.617 | 94.9 |
| Allocation Ratio as Covariate | -0.006 | 0.617 | 94.9 |
| Stratified by Site | -0.006 | 0.622 | 94.7 |
| Stratified by Allocation Ratio | -0.006 | 0.617 | 94.9 |
| Random Effect for Site | -0.828 | 0.611 | 72.3 |

<sup>†</sup>Time-to-event outcome results are based on Cox regression analyses of 10000 simulated randomized trials with 50% of patients randomized in sites using 1:1 allocation ratio and 50% randomized in sites using 1:3 allocation ratio. Randomization was stratified by study site. Total sample size for each trial was 4800 patients (1800 in the intervention group, 3000 in the control group). True hazard ratio 1.0. 95% confidence interval coverage is based on Wald-type confidence intervals. Baseline hazard was increased by 50% for sites randomized in a 1:3 ratio compared to sites randomized in a 1:1 ratio to produce bias in the unadjusted estimates.
<sup>‡</sup>Binary outcome results are based on logistic regression analyses of 10000 simulated randomized trials with 50% of patients randomized in sites using 1:1 allocation ratio and 50% randomized in sites using 1:3 allocation ratio. Randomization was stratified by study site. Total sample size for each trial was 4800 patients (1800 in the intervention group, 3000 in the control group). True odds ratio is 1.0. The analysis stratified by site used conditional logistic regression. Baseline probability of events was increased by 50% for sites randomized in a 1:3 ratio compared to sites randomized in a 1:1 ratio to produce bias in the unadjusted estimates.
<sup>§</sup>Continuous outcome results are based on linear regression analyses of 10000 simulated randomized trials with 50% of patients randomized in sites using 1:1 allocation ratio and 50% randomized in sites using 1:3 allocation ratio. Randomization was stratified by study site. Total sample size for each trial was 4800 patients (1800 in the intervention group, 3000 in the control group). True between-group difference is 0 units. Baseline mean was increased by 10 units (10%) for sites randomized in a 1:3 ratio compared to sites randomized in a 1:1 ratio to produce bias in the unadjusted estimates.

## 4.0 DISCUSSION

The allocation ratio can be varied by study site to maximize recruitment capacity and overcome asymmetric site-level constraints, such as limits on the ability to deliver one of the study interventions. Although the sample size requirements increase with use of ratios other than 1:1, recruitment capacity can increase to a greater extent for the right trial. This is most relevant in trials where many patients are eligible and willing to participate, and where the marginal cost of participant recruitment and follow-up is low. These conditions are common in many pragmatic trials which tend to have broad eligibility criteria, high acceptability to patients and providers, limited study procedures, and use existing databases for follow-up.

If a trial uses site-variable allocation ratios, its analysis must account for this variation or prognostic differences between sites using different ratios may bias the estimate of treatment effect. Simple covariate adjustment or stratification by site or allocation ratio addresses this issue. There may exist more complex scenarios not explored here.

If variable allocation (or any unequal allocation approach) is used, it should be clearly justified in the study protocol.[5] The sample size implications of constant unequal allocation are well-described,[6–9] but the treatment of variable allocation ratios in these texts focuses on adaptive techniques that either strive to balance groups or adapt based on treatment response, and introduce issues unrelated to our topic. Interested readers may explore statistical simulation studies that provide a deeper treatment of unequal allocation in randomized trials and evaluate complex randomization designs,[10] or focus on non-inferiority designs and allocation to more than two groups.[11]

The use of multiple allocation ratios that vary across study sites has received minimal attention. Douglas Altman, referencing earlier work by Armitage and Borchgrevink,[12] briefly discussed scenarios of varying allocation ratios.[13] He emphasized the need to analyze data from patients randomized in the same ratio separately and then pool across different ratios using meta-analysis (i.e., a stratified analysis as demonstrated here).

A review of methodological issues related to adaptive trials that change allocation ratios or add treatment arms during the trial—a unique form of change in allocation ratio—confirmed the scarce treatment of these issues.[14] Methods explored were predominantly concerned with type 1 error probability when adding a study arm and include simply pooling all data across different stages (before and after the addition of an arm), adjusting for design changes in a linear model, or apply adaptive methodologies that calculating P values separately for each stage and then combining them using principles derived from combination tests (the latter relevant to control type 1 error when design changes are driven by comparative interim analyses).[15] The authors conclude that adjusting for the design change in a linear model alone is acceptable if there have been no internal looks at the data to drive the design change and inflate type 1 error.[15]

This exploration of the site-variable allocation ratio was intended to be illustrative. The simulated scenarios address the core issues but lacked the complexities of real data. Stratifying by site or adjusting for site as a model covariate may not be feasible when there are many sites but few events; stratifying or adjusting for allocation ratio alone may be a more practical strategy in that scenario, potentially with a random effect for study site. The analyses focused on recruitment constraints induced by asymmetric limitations in intervention (or other trial procedure) delivery. In practice, sites may instead be limited by the number of eligible and interested patients, resources to deliver the comparator intervention, or resources to recruit and follow participants. Further, the presentation of patient characteristics and outcomes by study group at the end of the trial may feel unfamiliar to most readers and will require special attention.

Constraints on recruitment within study sites can be offset by significantly increasing the number of study sites, but such a strategy is always costly and sometimes completely infeasible. In some trials, researchers may be more selective with study sites and limit enrollment to sites with greater capacity to deliver the intervention. However, this may compromise generalizability of results to standard practice settings. Further, sites with less intervention capacity may serve a disadvantaged patient population and excluding them would limit access to research participation for patients in those communities. Researchers may instead consider tailoring the allocation ratio to sites’ constraints.

It is typical that investigators do not know all of the sites that will participate in the trial before the trial initiates recruitment. Therefore, it is difficult to predict the mix of sites that will use the different allocation ratios. If the constraints on intervention delivery are expected to be common, then investigators may wish to assume, for sample size calculations, that most or all sites will use the same unbalanced ratio. As sites that have little constraint on the intervention delivery enter the trial, randomizing patients in a 1:1 ratio will increase power and may allow investigators to readjust their sample size targets.

## 5.0 CONCLUSIONS

Site-variable allocation ratios can facilitate recruitment into randomized trials where logistics restrict enrolment asymmetrically (e.g., into one study group compared to others) in some—but not all—sites. Although the sample size requirements increase with use of ratios other than 1:1, recruitment capacity can increase to a greater extent for the right trial. Tailoring the allocation ratio to study sites’ operational constraints can optimize the combination of statistical efficiency and recruitment, but the analysis must adjust for the study site or allocation ratio.

## Acknowledgments

I thank Dr Amit X Garg for suggesting that I consider unequal allocation for a trial design of a hemodialysis filter. This became the impetus for the current investigation. I thank Dr Guangyong Zou for his generosity of time and wisdom in biostatistics, including with my questions around analytic methods in the variable allocation problem.

## Author Contributions

Dr Roshanov conceived the idea for this paper, designed and conducted the analyses, and wrote the manuscript.

## Data Availability

All data were synthetic; the statistical code to generate it is available upon request form the author.

